# Seroepidemiology among Employees of New York City Health and Hospitals during the First Wave of the SARS-CoV-2 Epidemic

**DOI:** 10.1101/2021.04.12.21255344

**Authors:** Alexander D. Bryan, Kathleen Tatem, Jillian Diuguid-Gerber, Caroline Cooke, Anya Romanoff, Nandini Choudhury, Michael Scanlon, Preeti Kishore, Elana Sydney, Joseph Masci, Parampreet Bakshi, Sahithi Pemmasani, Nichola J. Davis, Duncan Maru

## Abstract

**Objective:** Estimate the seroprevalence of SARS-CoV-2 antibodies among New York City Health + Hospitals healthcare workers, and identify demographic and occupational factors associated with SARS-CoV-2 antibodies among healthcare workers.

**Methods:** This was an observational, cross-sectional study using data from SARS-CoV-2 serological tests accompanied by a demographic and occupational survey administered to healthcare workers. Participants were employed by New York City Health + Hospitals (NYC H+H) and either completed serologic testing at NYC H+H between April 30 and June 30, 2020, or completed SARS-CoV-2 antibody testing outside of NYC H+H and were able to self-report results.

**Results:** Seven hundred twenty-seven survey respondents were included in analysis. Participants had a mean age of 46 years (SD= 12.19) and 543 (75%) were women. Two hundred fourteen (29%) participants tested positive or reported testing positive for the presence of SARS-CoV-2 antibodies (IgG+). Characteristics associated with positive SARS-CoV-2 serostatus were Black race (25% IgG+ vs. 15% IgG-, p=0.001), having someone in the household with COVID symptoms (49% IgG+ vs. 21% IgG-, p<0.001), or having a confirmed COVID-19 case in the household (25% IgG+ vs 5% IgG-, p<0.001). Characteristics associated with negative SARS-CoV-2 serostatus included working on a COVID patient floor (27% IgG+ vs. 36% IgG-, p=0.02), working in the ICU (20% IgG+ vs. 28% IgG-, p=0.03), or having close contact with a patient with COVID-19 (51% IgG+ vs. 62% IgG-, p=0.03).

**Conclusions:** Results underscore the significance of community factors and inequities might have on SARS-CoV-2 exposure for healthcare workers.

**What is already known about this subject?:** Healthcare workers are at risk of occupational transmission of SARS-CoV-2, and the risk of infection varies by demographic characteristics and work location.

**What are the new findings?:** Healthcare worker race and household contacts were significantly associated with SARS-CoV-2 seropositivity, while working on a COVID patient floor or ICU was associated with seronegativity.

**How might this impact on policy or clinical practice in the foreseeable future?:** Results underscore the significance of community factors and inequities on healthcare worker exposure to SARS-CoV-2, and the need to address these inequities at the community level where healthcare workers live.

## BACKGROUND

By the end of 2020 there were almost 85 million confirmed cases of SARS-CoV-2 and over 1.8 million deaths globally.^1^ The pandemic has placed enormous strains on healthcare systems and healthcare workers (HCW), including inpatient and community-based care providers as well as hospital administrators and support staff. As shown during prior infectious disease outbreaks, protecting HCW through adequate infection control and access to personal protective equipment (PPE) alongside general public health and preventative measures is critical to global pandemic response.^2^ In many countries, however, the current COVID-19 pandemic has led to truly unprecedented conditions for HCW and their physical and mental well-being.^3^

Epidemiologic and serologic data on SARS-CoV-2 among HCW are essential to guide health care systems and public health policies and protect HCW.^4^ Early data from China suggested that HCW were at high risk of SARS-CoV-2 infection,^5^ and since then a significant body of literature has emerged on SARS-CoV-2 among HCW.^6 7^ A systematic review and meta-analysis of 97 studies including 230,398 HCW found a pooled SARS-CoV-2 prevalence rate of 11% in studies using rtPCR tests and 7% using serum antibody tests, but there were insufficient data in most studies to assess risk factors and exposure levels.^7^ HCW are at risk of occupational transmission of SARS-CoV-2 in inpatient and outpatient settings, particularly with inadequate PPE or infection control procedures. ^8 9^ HCW are also at risk for community transmission of SARS-CoV-2,^10^ while household members of HCW may be at higher risk compared to the general public.^11^

Among HCW, the risk of infection varies by demographic characteristics, cadre of HCW, and work location, with systemic racism playing a clear role in inequities.^12^ Additionally, among HCW with a job setting reported, most infections were associated with nursing and residential care facilities (67%) compared to hospital settings (18%). There are few data on SARS-CoV-2 among community-based HCW and other social service workers who may have different demographic and occupational risk profiles.^13^ Approximately six weeks into the pandemic, the largest public hospital system in the US, New York City Health and Hospitals (NYC H+H), initiated universal, voluntary serologic testing among all employees. We invited employees who were undergoing serologic testing to participate in a survey to assess demographic and occupational factors associated with serostatus. Specifically, we aimed to: (1) estimate the seroprevalence of SARS-CoV-2 antibodies among NYC H+H HCW, and (2) identify demographic and occupational factors associated with SARS-CoV-2 antibodies among NYC H+H HCW.

## METHODS

### Study Setting

New York City H+H employs over 40,000 people in a wide range of clinical and non-clinical positions at 11 acute care hospitals and more than 70 community facilities across the city’s five boroughs. This study leveraged universal, voluntary SARS-CoV-2 antibody testing which was available at NYC H+H starting in April 2020. Testing was open to all employees of NYC H+H and was available at NYC H+H ambulatory settings across the city.

### Study Design

This was an observational, cross-sectional study using data from SARS-CoV-2 serological tests accompanied by a demographic and occupational survey administered to HCW at NYC H+H. The primary endpoint was SARS-CoV-2 serological testing outcome, stratified by key demographic and occupational characteristics reported through the survey. This study was approved by the Institutional Review Board of the Biomedical Research Alliance of New York

To assess the risk of contracting Covid-19, we developed a modified survey based on the World Health Organization’s *Protocol for assessment of potential risk factors for 2019-novel coronavirus (2019-nCoV) infection among health care workers in a health care setting*.^4^ Surveys were self-administered in paper form at testing sites or electronically through a REDCap survey online. Survey respondents who completed SARS-CoV-2 antibody testing at NYC H+H were matched to serological results in the NYC H+H electronic health record. Results of SARS-CoV-2 rtPCR testing were included in secondary analyses when available.

### Study Population

Surveys were self-administered in paper form at testing sites, and an electronic version of the survey was emailed system-wide to all NYC H+H employees. All NYC H+H employees who had serologic testing from April 30 to June 30, 2020 were invited to participate. Additionally, employees who received antibody testing outside of NYC H+H between April 30 and June 30, 2020 could complete the survey and self-report serology results.

In order to be eligible for the study, participants needed to meet the following criteria:

1. Employed by NYC H+H and either A) completed serologic testing at NYC H+H or B) completed SARS-CoV-2 antibody testing outside of NYC H+H and were able to self-report results;
2. 18 years of age or older;
3. Capable of providing consent to participate in the study, including English or Spanish fluency.

Additionally, limited data on demographics and seropositivity is included on all H+H employees who completed antibody testing from April 30 to June 30, 2020 but did not participate in the survey study.

### Key Definitions

#### SARS-CoV-2 antibody status

SARS-Cov-2 antibody status was assigned based on serological results from the NYC H+H EHR. Serological testing at NYC H+H was performed using the SARS-CoV-2 IgG Assay from Abbott Laboratories Inc. For participants who completed serologic testing at an outside institution, self-report results were used.

#### Demographic Characteristics

Demographic characteristics were self-reported. Sex was captured as male or female. Age was calculated from date of birth, and country of origin analyzed as US born or non-US born. Participant zip code of primary residence was used to determine borough (i.e. Manhattan, Brooklyn, Bronx, Queens, Staten Island, non-NYC). Race and ethnicity were reported separately and variables were combined together into one variable with mutually exclusive categories for analysis, with Black representing non-Hispanic Black participants and White representing non-Hispanic White participants.

#### Occupation

Participants selected occupation from a pre-specified list or selected ‘Other’ and provided a free text response answer. Free text response answers were re-grouped into existing or new categories when appropriate. The following additional groupings emerged: care coordination, pharmacy, and counseling (distinct from social worker).

### Statistical Analysis

We used descriptive statistics to explore demographic characteristics, case exposure, occupational setting and SARS-CoV-2 serological test results of the study population, and explored the association of serological test results with demographic (e.g., zip code) and occupational characteristics and exposures. Continuous measures were expressed as means and standard deviation, categorical variables as counts and proportions. We conducted bivariate analyses using Fisher’s exact test for categorical variables and t-tests, as appropriate, with significance set at α=0.05. In addition, subsequent chi-square tests were conducted for age using 21-34 as a reference group and for race using White as a reference group. Missing values were removed for all calculations resulting in differing denominators for each demographic question and exposure-related variable. R Studio Version 1.3.1093 using the tidyverse and gmodels packages was used for statistical analysis.

## RESULTS

In total, 19,107 staff completed antibody testing at NYC H+H from April 30 to June 30. During that time, there were 1,671 respondents to the demographic and occupational survey, and 727 survey respondents were matched to SARS-CoV-2 antibody results, either through the NYC H+H Electronic Health Record (EHR) or through self-report. Reasons for being unable to match survey respondents to serologic results included surveys missing full name or date of birth (n=766), or name and date of birth match not found in EHR (n=178).

Participants had a mean age of 45.97 years (SD= 12.19) and 543 (75%) were women. Two-hundred and sixty-four (36%) self-reported as White, 146 (20%) as Hispanic, 128 (18%) as Black, 148 (20%) as Asian, 19 (2.6%) as Other, 11 (1.5%) as Multiracial, 4 (0.6%) as Pacific Islander, and 6 (0.8%) as missing. Of the 705 participants who reported their country of origin, 322 (46%) were US-born. Over 70 different countries of origin were reported. A total of 539 (75%) participants reported living in New York City. Over half of the sample, 407 (58%) participants, reported having known close contact with at least 1 SARS-CoV-2 positive patient, 254 (37%) noted close contact with materials of at least 1 SARS-CoV-2 positive patient, 210 (29%) reported living with someone experiencing COVID-19 symptoms, and 76 (11%) reported living with someone with confirmed COVID-19.

Supplemental eFigure 1A-C compares the study participants to the 19,107 H+H staff who completed serology testing between April 30 and June 30, 2020, and the overall H+H workforce (estimated 42,000). The three groups were similar in sex (female 75% survey sample vs. 70% tested workforce vs. 69% total workforce). Race/ethnicity differed between the study participants, H+H staff completing serology testing, and the total H+H workforce, with study participants over-representing White employees and under-representing Black employees.

Of the 727 participants included in the study, 658 (91%) participants completed an antibody test directly at an H+H site, and 69 (9.5%) participants reported antibody tests from an outside location. Overall, 214 (29%) participants tested positive or reported testing positive for the presence of SARS-CoV-2 antibodies. Comparatively, of the 19,107 H+H staff who completed serology testing between April 30 and June 30, 4,610 (24%) tested positive for the presence of SARS-CoV-2 antibodies. Supplemental eFigure2A-C shows seropositivity of survey participants and 19,107 H+H staff by sex, race/ethnicity and age.

Table 1 reports key demographic factors and community and occupational exposures according to SARS-CoV-2 antibody status (SARS-CoV-2 IgG positive or negative). Both groups were majority female (378 IgG- [75%] vs.164 IgG+ [78%]), similar in age (mean [SD], 45 [12] years IgG- vs. 46 [11] years IgG+) (eTable1), and majority born in the United States (301 IgG- [61%] vs. 121 IgG+ [58%]). Compared to those who were negative for SARS-CoV-2 antibodies, a larger percentage of respondents positive for SARS-CoV-2 antibodies were Black (54 IgG+ [25%] vs. 74 IgG- [15%]) (Table 2), had someone in their household with COVID symptoms (104 IgG+ [49%] vs 106 IgG- [21%]), and had a COVID-19 confirmed case in their household (52 IgG+ [25%] vs 24 IgG- [5%]). A lower percentage of respondents positive for SARS-CoV-2 antibodies worked on a COVID patient floor (58 IgG+ [27%] vs.185 IgG- [36%]), in the Intensive Care Unit (ICU) (43 IgG+ [20%] vs.143 IgG- [28%]), or had close contact with a patient with COVID-19 (106 IgG+ [51%]) vs. 301 IgG- [62%]).

**Table 1:**
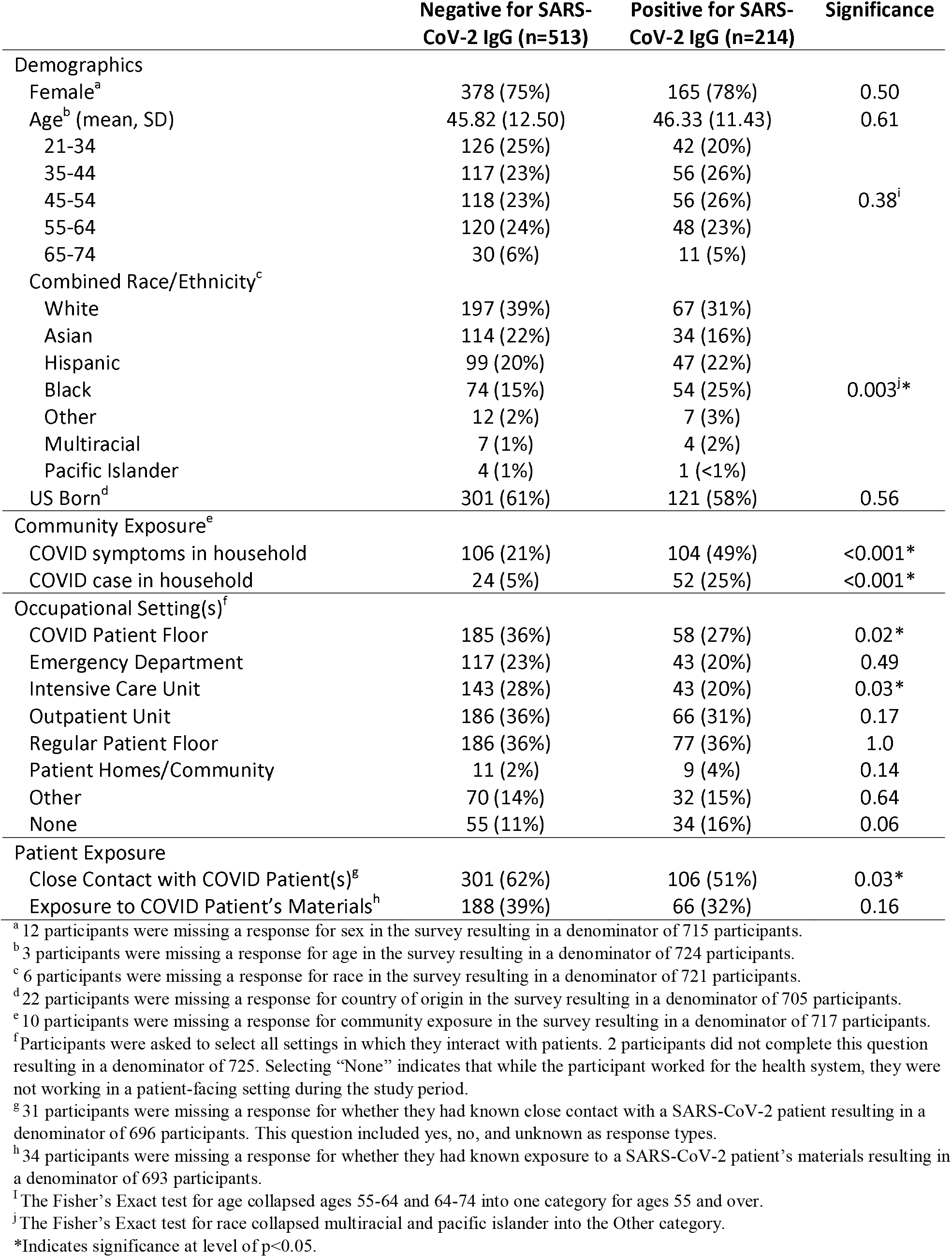
Demographics, case exposure, and occupational setting by SARS CoV-2 antibody status.

**Table 2.**
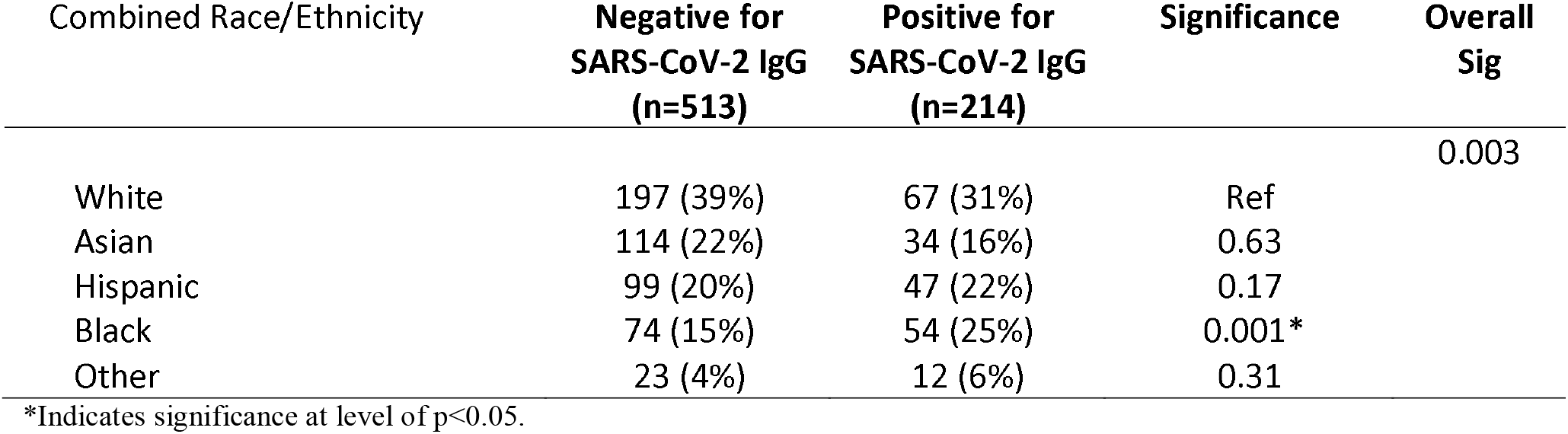
Exploratory Post-Hoc Analyses of combined Race/Ethnicity by SARS CoV-2 antibody status.

| Combined Race/Ethnicity | Negative for<br>SARS-CoV-2 IgG<br>(n=513) | Positive for<br>SARS-CoV-2 IgG<br>(n=214) | Significance | Overall<br>Sig |
| --- | --- | --- | --- | --- |
| White | 197 (39%) | 67 (31%) | Ref | 0.003 |
| Asian | 114 (22%) | 34 (16%) | 0.63 |  |
| Hispanic | 99 (20%) | 47 (22%) | 0.17 |  |
| Black | 74 (15%) | 54 (25%) | 0.001* |  |
| Other | 23 (4%) | 12 (6%) | 0.31 |  |

\*Indicates significance at level of $p < 0.05$ .

Figure 1 shows the seropositivity rate for SARS-CoV-2 IgG by self-reported staff occupation. There were 209 (28%) Doctors, Nurse Practitioners (NP) or Physician Assistants (PA), 154 (21%) Registered Nurses (RN), 87 (12%) Administrators 15 (2.1%) Care Coordinators, 15 (2.1%) Pharmacy staff, 12 (1.7%) Radiology staff, and 8 (1.1%) in Food services. Crude seropositivity rates for Doctors, NPs and PAs (34 IgG+) was 16%, compared to 34% for RNs (52 IgG+), 39% for Administrators (34 IgG+), and 62% for Food services (5 IgG+).

**Figure 1.**
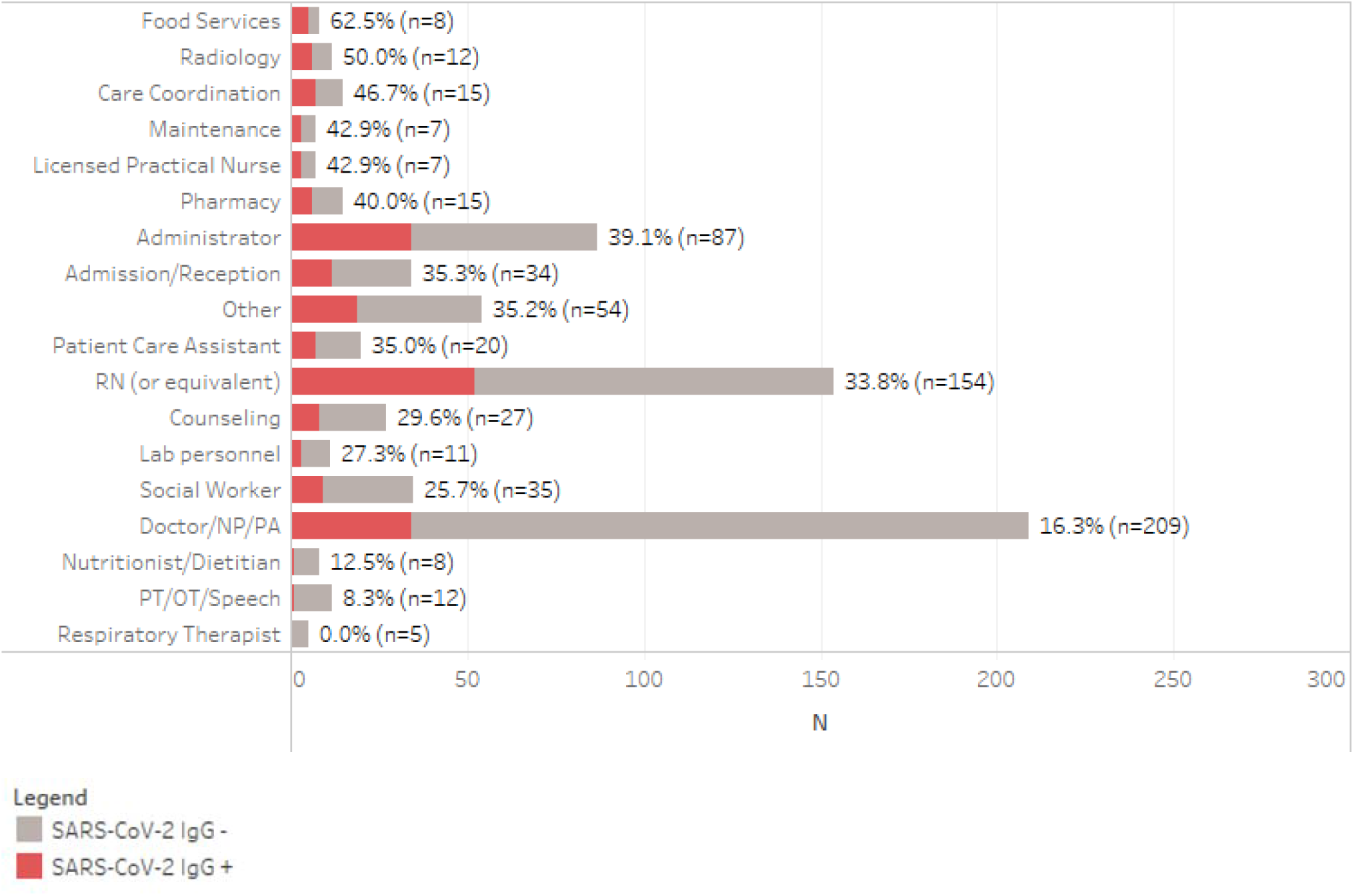
Percent seropositive for SARS-CoV-2-IgG by reported Staff Occupation (n=720)

Figure 2 shows seropositivity rates of SARS-CoV-2 IgG by how frequently PPE was used when indicated, and Figure 3 shows reported PPE availability within the healthcare facility. The majority of respondents reported ‘Always’ or ‘Most of the time’ wearing a medical or surgical mask (681 respondents [98%]) when indicated, with 627 (92.6%) reporting that they were available ‘Always’ or ‘Most of the time’. A total of 430 respondents reported ‘Always’ or ‘Most of the time’ (72%) wearing a respiratory mask (e.g. N95), with 169 (28%) ‘Never’, ‘Rarely’, or ‘Occasionally’ wearing the respiratory mask when indicated. The availability of the respiratory masks varied, with 180 (30%) saying it was ‘Always’ available, and 235 (39%) reporting them available ‘Most of the time’. The crude seropositivity rates among the 169 that ‘Never’, ‘Rarely’, or ‘Occasionally’ wore respiratory masks was 36.7%, compared to 26.3% for the 430 that wore a respiratory mask ‘Always’ or ‘Most of the time’. For all other PPE, the majority of respondents ‘Always’ wore PPE when indicated, except for impermeable gowns, coverall/body suits, shoe covers and HEPA filters on endotracheal tube for intubated patients. Availability of PPE varied by PPE type, with most being available ‘Always’ or ‘Most of the time,’ except Coverall/body-suits.

**Figure 2.**
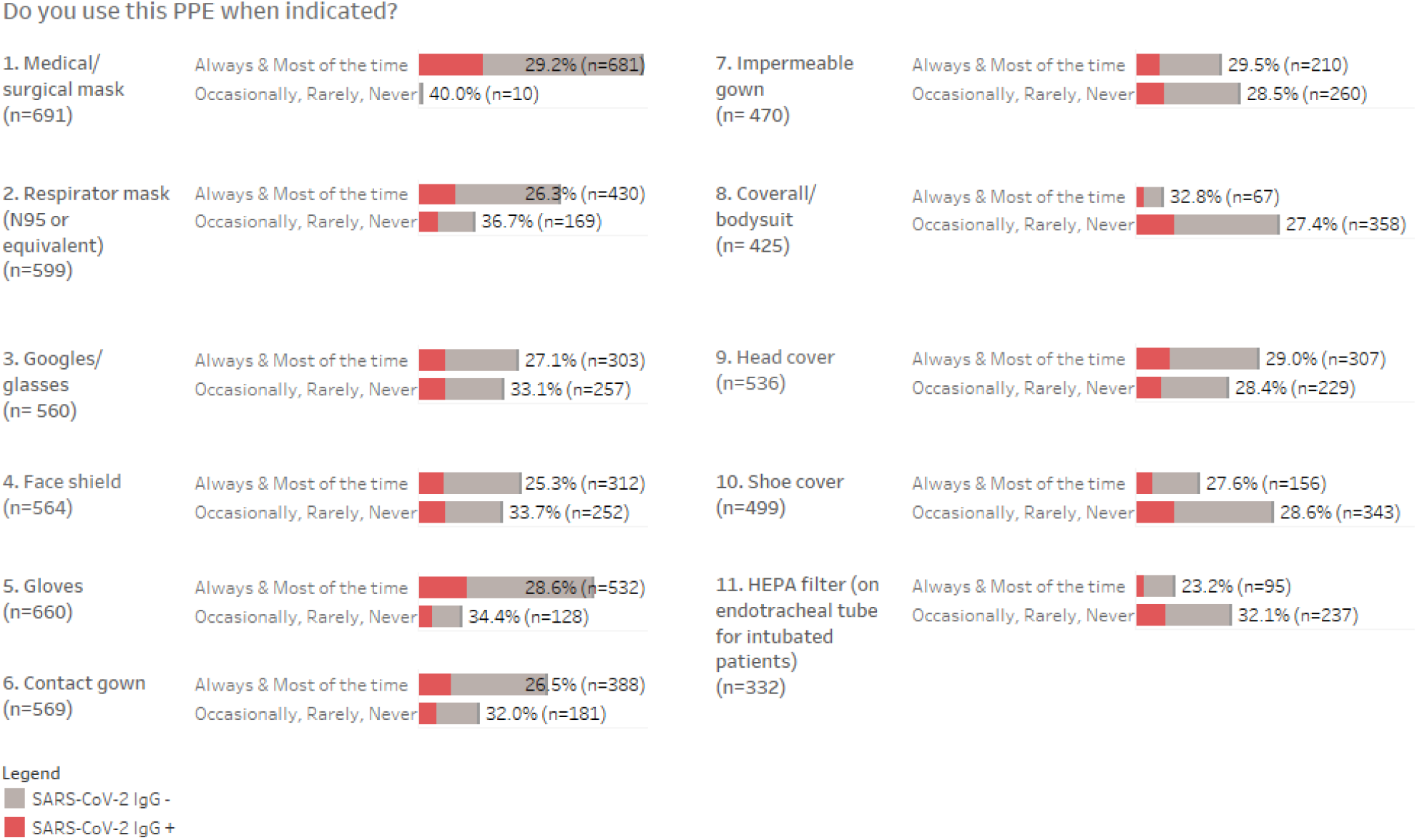
Percent seropositivity for SARS-CoV-2-IgG by how frequently PPE was used when indicated. Excludes patients who noted PPE type was Not Applicable or missing.

**Figure 3.**
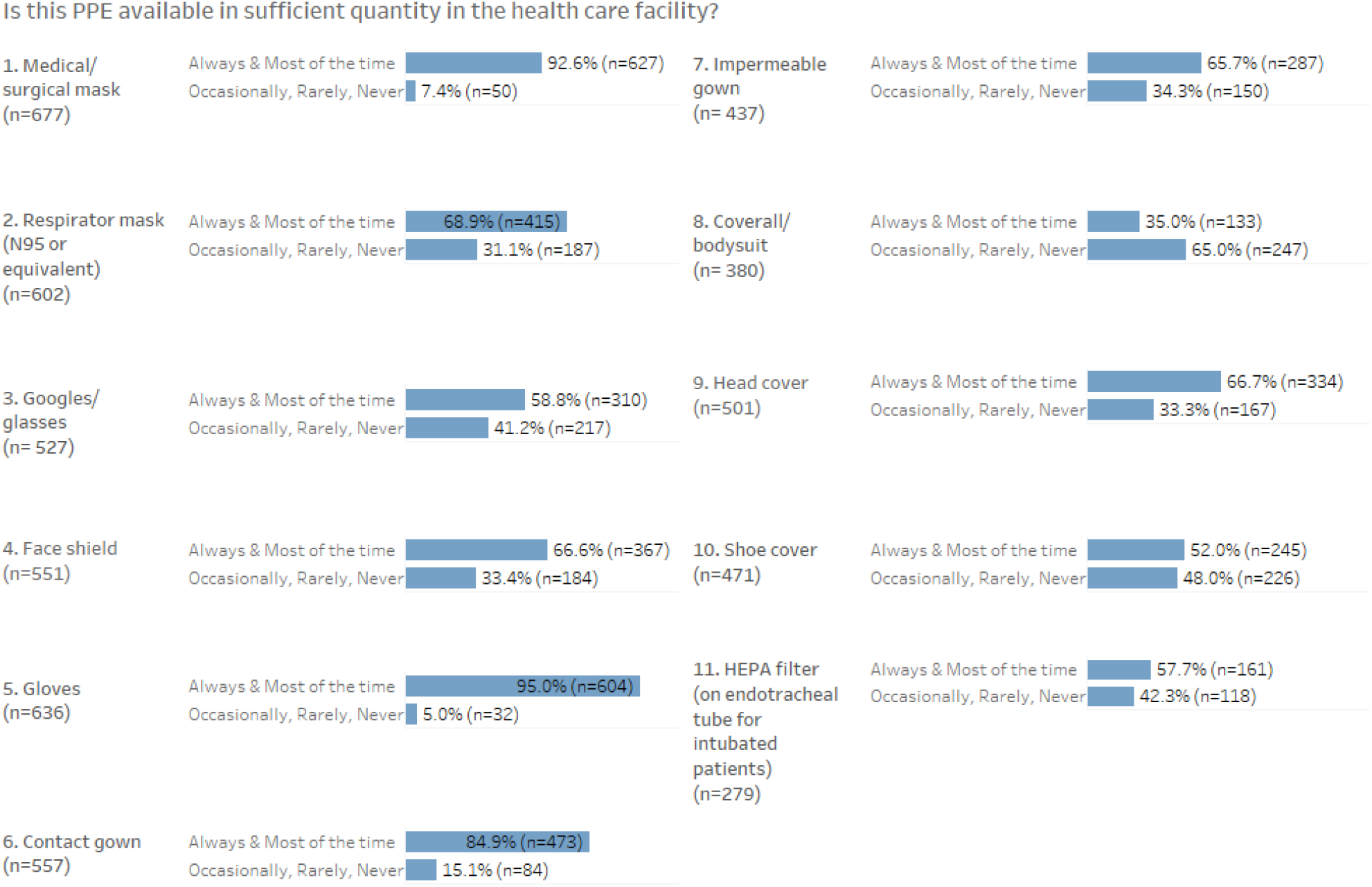
PPE availability. Excludes patients who noted PPE type was Not Applicable or missing.

Supplementary eTable 2 shows the distribution of seropositive study participants by borough of primary residence compared to the distribution of cumulative COVID cases in New York City by borough from February 29 to June 30, 2020. The distribution of seropositive study participants living in NYC was highest in Queens (34%), followed by Brooklyn (25%) and the Bronx (20%). The distribution of cumulative cases reported in NYC was also highest in Queens (30%), followed by Brooklyn (28%) and the Bronx (22%).

## DISCUSSION

In this system-wide survey of employees during the first wave of the COVID-19 epidemic in New York City, 29% of study participants tested positive for SARS-CoV-2 antibodies. This was similar to the overall seropositivity of employees tested at NYC H+H, and among the highest rates of employee seropositivity reported by health systems in the NYC area during this period.^14 15^ One possible difference to explain these higher rates of seropositivity at NYC H+H compared to rates reported from health systems in Manhattan and Long Island could be the higher rates of community exposure at the time in Queens, Brooklyn and the Bronx, where the majority of NYC H+H employees live. Study participant seropositivity was highest for employees living in Queens, Brooklyn and the Bronx, while the cumulative distribution of cases in New York City at the time closely followed this trend. Employee seropositivity may have been largely driven by community spread at this phase of the pandemic in NYC, as seropositivity was most strongly associated with household COVID-19 contact. However, a lack of temporal data limits whether this might be interpreted as community transmission to HCW or HCW transmitting to household members.

Doctor/NP/PA were the most frequently reported occupations among respondents, and they were among the lowest in seropositivity. This may reflect differing patient exposures or differing use of PPE, as RNs (or equivalent) were the second largest occupational group surveyed and had over double the seropositivity rate. Seropositivity rates were similarly higher among other large occupational groups surveyed such as Administrator and Admission/Reception, which is notable as these are patient-facing but typically non-clinical occupation groups. In fact, working on a COVID-19 patient floor or ICU was associated with testing negative for SARS-CoV-2 antibodies, as was close contact with COVID-19 patients. These results potentially indicate greater adherence to PPE, greater availability of PPE, or clearer recommendations on PPE usage for providers working in those settings, which offset patient exposure.

In examining employee use of PPE, consistent use of certain equipment was associated with lower reported rates of seropositivity. Most notably, employees who reported using an N95 mask ‘Always’ or ‘Most of the time’ when indicated had a seropositivity rate of 26.3%, while employees using an N95 ‘Occasionally’, ‘Rarely’ or ‘Never’ had seropositivity rates of 36.7%. Although a majority (58%) of respondents reported always wearing an N95 when indicated, only 30% reported an N95 was always available in sufficient quantity in the health care facility. This not only reflects the low supply of PPE in NYC at the height of the pandemic, but also the shortfall in PPE that HCW needed to bridge by stretching available supplies. Practices to stretch PPE supply reported elsewhere in the literature included extended use, reuse, or decontamination procedures, and may account for this disparity in responses.^16 17^

Black race was strongly associated with employee seropositivity, echoing broader racial inequities seen in the community during the COVID-19 pandemic.^18 19^ In the US, among 100,570 cases of SARS-CoV-2 and 641 deaths among HCW up to July 16, 2020 that were reported to the Centers for Disease Control and Prevention, Asian and Black HCW were over-represented among fatal cases, and health care support workers, nurses, and administrative staff were the most common occupational types among those infected.^20^ In a prospective cohort study using self-reported data through a smartphone application among almost 100,000 HCW in the UK and US, Asian, Black, and other minority ethnic HCW were at higher risk of SARS-CoV-2 infection and more likely to report having inadequate access to PPE compared to White HCW.^21^ These results underscore the connection between HCW and their communities with regards to these systemic inequities, and highlight the continued need to urgently address these disparities for the wellbeing of patients as well as the workers caring for those patients.

There were several limitations to our study. First, convenience sampling was used to enroll study participants, and while we recruited study participants from a voluntary and universal screening program, there was a potential for selection bias in terms of occupation type and level of exposure in survey respondents. In addition, there may have been socioeconomic factors related to participation that could be strongly tied to demographic and occupational characteristics. We attempted to partially mitigate these limitations by comparing aggregate participant demographics with all employees undergoing antibody testing, as well as with overall NYC H+H employee demographics. This revealed under-representation of certain groups in the study participants, most notably Black employees. Given that Black employees were also found to be more likely to have SARS-CoV-2 antibodies, this may have skewed overall positivity rates and excluded differing exposure factors. We were also unable to determine when employees with positive SARS-CoV-2 antibodies were infected. And with continuously changing guidelines around PPE during the initial surge, it is difficult to link exposure and infection with evolving PPE practices. Furthermore, employees who were sick or were suspected to have SARS-CoV-2 may have been less likely to get serological testing, and participants who previously tested PCR positive for SARS-CoV-2 may or may not have chosen serological testing. Our study did not account for HCW who were currently hospitalized or had died. These factors may have further biased our sample and underestimated the burden of SARS-CoV-2 infection in this population. Finally, this was a descriptive analysis and it is possible that some findings may be due uncontrolled confounding factors.

## CONCLUSION

Employees at a large, public hospital system reported a seropositivity rate of 29% during the first wave of the COVID-19 epidemic in New York City. This was among the highest employee seropositivity rates reported in NYC, and the risk of exposure varied significantly by employee demographics, occupation, and work location. Results underscore the need to address exposure risks for HCW across occupational settings, including appropriate PPE, as well as address broader inequities at the community level where HCW live.

## Supporting information

Survey

## Data Availability

Deidentified data are available upon reasonable request from the corresponding author.

## SUPPLEMENTARY MATERIALS

Study Survey

eFigures 1-2

eTables 1-2

