## Supplementary material for "Seroepidemiology among Employees of New York City Health and Hospitals during the First Wave of the SARS-CoV-2 Epidemic": Survey

**New York City Health + Hospitals Employee COVID-19 Antibodies Survey**

| | |
|---|---|
| Name | |
| Medical record number (if known) | |
| **1. Data collection** | |
| 1.1. Date of survey (DD/MM/YYYY) | ___/___/___ |
| 1.2. Format of survey | □ In person<br>□ Web-based<br>□ Telephone |
| 1.3. Permission to contact for future research?<br>"We are still learning about the meaning of these antibody results and we may follow participants over time to find out whether they have gotten sick or to repeat antibody testing. Is it okay if we contact you in the future to participate in another study?" | □ Yes □ No<br>*If no, skip to section 2 |
| 1.4. Primary phone number | |
| 1.5. Secondary phone number | |
| 1.6. Email address | |
| **2. Demographic Information** | |
| 2.1. Date of birth (DD/MM/YYYY) | __/__/__ |
| 2.2. Sex | □ Male □ Female |
| 2.3 Zip code of primary residence | |
| 2.4. Country of birth | |
| 2.5. Race | □ American Indian<br>□ Asian<br>□ Black/African American<br>□ Pacific Islander<br>□ White<br>□ Other ________________________ |
| 2.6. Ethnicity | □ Hispanic/Latino<br>□ Not Hispanic/Latino |
| 2.7. Primary work location | □ Bellevue Hospital<br>□ Coney Island Hospital<br>□ Elmhurst Hospital<br>□ Harlem Hospital<br>□ Jacobi Hospital<br>□ Kings County Hospital<br>□ Lincoln Hospital<br>□ Metropolitan Hospital<br>□ North Central Bronx Hospital<br>□ Queens Hospital<br>□ Woodhull Hospital<br>□ Belvis |

BRANY
Version Date: 05/29/2020

| | |
|---|---|
| | □ East New York<br>□ Gouverneur<br>□ Morrisania<br>□ Sydenham<br>□ Vanderbilt<br>□ Gotham Health- Neighborhood Health Clinic<br>□ Carter<br>□ Coler<br>□ McKinney<br>□ Seaview<br>□ Home and Community Based (ie Community Care)<br>□ Central Office |
| 2.8. Occupation in health care facility | □ Admission/reception clerks<br>□ Food services staff<br>□ Healthcare administrator<br>□ Lab personnel<br>□ Licensed practical nurse (or equivalent)<br>□ Maintenance/cleaning staff<br>□ Medical doctor/ nurse practitioner/ physician assistant<br>□ Nutritionist/dietitian<br>□ Patient care assistant<br>□ Patient transporter<br>□ Phlebotomist<br>□ Physical /occupational/ speech therapist<br>□ Radiology / x-ray technician<br>□ Registered nurse (or equivalent)<br>□ Respiratory therapist<br>□ Social Worker<br>□ Other________________ |
| 2.9. During the COVID pandemic, are you performing health care duties outside of your usual responsibilities? | □ Yes □ No |
| 2.10. In what setting(s) do you interact with patients? | □ Regular patient floor<br>□ COVID patient floor<br>□ Intensive care unit<br>□ Emergency department<br>□ Outpatient Clinic<br>□ Patient homes and communities<br>□ Other________________<br>□ None |

BRANY
Version Date:
05/29/2020

| **3. Prior COVID-19 testing/symptoms** | |
|---|---|
| 3.1. Did you receive nasal or nasopharyngeal swab testing for COVID-19 before? | □ Yes □ No |
| 3.2. If so, what was the result? | □ Positive □ Negative □ N/a |
| 3.3 Permission to access participant records for SARS-CoV-2 PCR results? "Some employees have been tested for coronavirus via nasopharyngeal swab PCR testing. Employees may or may not want to share their past test results. Do you allow us to access your COVID-test PCR results from our computer system?" | □ Yes □ No |
| 3.4 Did you receive COVID-19 antibody blood testing from a program other than the Health and Hospitals employee testing program, such as through Mount Sinai, the Department of Health, the CDC, or an outside group such as CityMD? Y/N | □ Yes □ No |
| 3.5 If so, what was the result? | □ Positive (you have antibodies to COVID-19)<br>□ Negative (you do not have antibodies to COVID-19)<br>□ Indeterminate (the test couldn't tell if you have antibodies or not)<br>□ I don't know (you do not know your results) |
| 3.6. Have you experienced any of the following symptoms since January 1, 2020? | |
| -Fever (greater than 100 degrees Fahrenheit) | □ Yes □ No |
| -Tiredness | □ Yes □ No |
| -Dry cough | □ Yes □ No |
| -Shortness of breath | □ Yes □ No |
| -Aches and pains | □ Yes □ No |
| -Sore throat | □ Yes □ No |
| -Runny nose | □ Yes □ No |
| -Diarrhea | □ Yes □ No |
| -Nausea | □ Yes □ No |
| -Decreased sense of smell or taste | □ Yes □ No |
| 3.7. Have you missed work because of any of the above symptoms since January 1, 2020? | □ Yes □ No |
| **4. Community exposure** | |
| 4.1. Has anyone living in your household had symptoms suggestive of COVID-19 (see above symptom list)? | □ Yes □ No |
| 4.2. Has anyone living in your household had a positive test for COVID-19? | □ Yes □ No |
| **5. Infection prevention and control measures** | |

| | |
|---|---|
| 5.1. When was your most recent infection prevention and control (IPC) training within the health care facility? | □ Within the last year<br>□ Greater than one year ago<br>□ Never<br>□ I don't remember |
| 5.2. How much cumulative IPC training (standard precautions, additional precautions, donning/doffing personal protective equipment) have you had since January 1, 2020? | □ None<br>□ Less than 1 hour<br>□ More than 1 hour |
| 5.3. Do you follow recommended hand hygiene practices? | □ Always<br>□ Most of the time<br>□ Occasionally<br>□ Rarely |

| 5.4. | **Do you this use PPE when indicated?** | **Is this PPE available in sufficient quantity in the health care facility?** |
|---|---|---|
| **Medical/surgical mask** | □ Always<br>□ Most of the time<br>□ Occasionally<br>□ Rarely<br>□ Never<br>□ Not applicable | □ Always<br>□ Most of the time<br>□ Occasionally<br>□ Rarely<br>□ Never<br>□ Not applicable |
| **Respirator mask (N95 or equivalent)** | □ Always<br>□ Most of the time<br>□ Occasionally<br>□ Rarely<br>□ Never<br>□ Not applicable | □ Always<br>□ Most of the time<br>□ Occasionally<br>□ Rarely<br>□ Never<br>□ Not applicable |
| **Goggles/glasses** | □ Always<br>□ Most of the time<br>□ Occasionally<br>□ Rarely<br>□ Never<br>□ Not applicable | □ Always<br>□ Most of the time<br>□ Occasionally<br>□ Rarely<br>□ Never<br>□ Not applicable |
| **Face shield** | □ Always<br>□ Most of the time<br>□ Occasionally<br>□ Rarely<br>□ Never<br>□ Not applicable | □ Always<br>□ Most of the time<br>□ Occasionally<br>□ Rarely<br>□ Never<br>□ Not applicable |
| **Gloves** | □ Always<br>□ Most of the time<br>□ Occasionally | □ Always<br>□ Most of the time<br>□ Occasionally |

BRANY
Version Date:
05/29/2020

| | | |
|---|---|---|
| | ☐ Rarely<br>☐ Never<br>☐ Not applicable | ☐ Rarely<br>☐ Never<br>☐ Not applicable |
| **Contact gown** | ☐ Always<br>☐ Most of the time<br>☐ Occasionally<br>☐ Rarely<br>☐ Never<br>☐ Not applicable | ☐ Always<br>☐ Most of the time<br>☐ Occasionally<br>☐ Rarely<br>☐ Never<br>☐ Not applicable |
| **Impermeable gown** | ☐ Always<br>☐ Most of the time<br>☐ Occasionally<br>☐ Rarely<br>☐ Never<br>☐ Not applicable | ☐ Always<br>☐ Most of the time<br>☐ Occasionally<br>☐ Rarely<br>☐ Never<br>☐ Not applicable |
| **Coverall/bodysuit** | ☐ Always<br>☐ Most of the time<br>☐ Occasionally<br>☐ Rarely<br>☐ Never<br>☐ Not applicable | ☐ Always<br>☐ Most of the time<br>☐ Occasionally<br>☐ Rarely<br>☐ Never<br>☐ Not applicable |
| **Head cover** | ☐ Always<br>☐ Most of the time<br>☐ Occasionally<br>☐ Rarely<br>☐ Never<br>☐ Not applicable | ☐ Always<br>☐ Most of the time<br>☐ Occasionally<br>☐ Rarely<br>☐ Never<br>☐ Not applicable |
| **Shoe covers** | ☐ Always<br>☐ Most of the time<br>☐ Occasionally<br>☐ Rarely<br>☐ Never<br>☐ Not applicable | ☐ Always<br>☐ Most of the time<br>☐ Occasionally<br>☐ Rarely<br>☐ Never<br>☐ Not applicable |
| **HEPA filter (on endotracheal tube for intubated patients)** | ☐ Always<br>☐ Most of the time<br>☐ Occasionally<br>☐ Rarely<br>☐ Never<br>☐ Not applicable | ☐ Always<br>☐ Most of the time<br>☐ Occasionally<br>☐ Rarely<br>☐ Never<br>☐ Not applicable |

### 6. Exposures to COVID-19 infected patients

| | |
|---|---|
| 6.1. Have you had close contact (within 6 feet/2 meters) with at least one COVID-19 positive patient since his/her admission? | □ Yes □ No □ Unknown<br>*If no, skip to section 7 |
| 6.2. If yes, for approximately how many encounters (total)? | □ 1 to 9<br>□ 10 to 19<br>□ 20 to 29<br>□ 30 to 39<br>□ 40 to 59<br>□ 50 or more |
| 6.3. For how long each time? | □ <5 minutes<br>□ 5-15 minutes<br>□ >15 minutes |
| 6.4. Did you wear a respirator mask (N95 or equivalent)? | □ Always<br>□ Most of the time<br>□ Occasionally<br>□ Rarely<br>□ Never |
| 6.5. If you were wearing a respirator, was it fit tested? | □ Yes □ No □ Unknown |
| 6.6. Have you needed to re-use a respirator mask (N95 or equivalent) for more than one day of patient care? | □ Yes □ No □ N/a |
| 6.7. If yes, for how many days do you use each mask on average? | ______ days |
| **7. Exposures to COVID-19 infected patient materials** | |
| 7.1. Have you had direct contact with a COVID-19 infected patient's materials since his/her admission? *(Patient's materials: personal belongings, linens, medical equipment, surfaces, etc. that the patient may have had contact with)* | □ Yes □ No □ Unknown<br>*If no, skip to section 8 |
| 7.2. If yes, which materials? | Check all that apply:<br>□ Clothes<br>□ Personal items<br>□ Linens<br>□ Medical devices used on the patient<br>□ Medical equipment connected to the patient (e.g. ventilator, infusion pump etc)<br>□ Bed<br>□ Bathroom<br>□ Bedside table<br>□ Other: ________________ |
| 7.3. Approximately how many times (total)? | □ None |

| | |
|---|---|
| | □ 1-4<br>□ 5-9<br>□ 10-19<br>□ 20 or more |

**8. Exposures to aerosolizing procedures**

| | **How many times have you been involved in the following procedures (in the patient room/at bedside)?** | **What proportion of the time were you wearing appropriate PPE?** |
|---|---|---|
| **Intubation** | □ None<br>□ 1-4<br>□ 5-9<br>□ 10-19<br>□ 20 or more | □ Always<br>□ Most of the time<br>□ Occasionally<br>□ Rarely<br>□ Never<br>□ Not applicable<br><br>What type? (Check all that apply):<br>□ Medical mask<br>□ Face shield<br>□ Gloves<br>□ Goggles/glasses<br>□ Contact gown<br>□ Impermeable gown<br>□ Coverall<br>□ Head cover<br>□ Respirator (e.g. N95 or equivalent)<br>□ Shoe covers |
| **Extubation** | □ None<br>□ 1-4<br>□ 5-9<br>□ 10-19<br>□ 20 or more | □ Always<br>□ Most of the time<br>□ Occasionally<br>□ Rarely<br>□ Never<br>□ Not applicable<br><br>What type? (Check all that apply):<br>□ Medical mask<br>□ Face shield<br>□ Gloves<br>□ Goggles/glasses<br>□ Contact gown<br>□ Impermeable gown<br>□ Coverall<br>□ Head cover<br>□ Respirator (e.g. N95 or equivalent)<br>□ Shoe covers |
| | **How many times have you been involved in the following procedures** | **What proportion of the [illegible] wearing appropriate PPE [illegible]** |

BRANY
Version Date:
05/29/2020

| | | |
|---|---|---|
| | **(in the patient room/at bedside)?** | |
| **Bronchoscopy** | □ None<br>□ 1-4<br>□ 5-9<br>□ 10-19<br>□ 20 or more | □ Always<br>□ Most of the time<br>□ Occasionally<br>□ Rarely<br>□ Never<br>□ Not applicable<br><br>What type? (Check all that apply):<br>□ Medical mask<br>□ Face shield<br>□ Gloves<br>□ Goggles/glasses<br>□ Contact gown<br>□ Impermeable gown<br>□ Coverall<br>□ Head cover<br>□ Respirator (e.g. N95 or equivalent)<br>□ Shoe covers |
| **Open suctioning** | □ None<br>□ 1-4<br>□ 5-9<br>□ 10-19<br>□ 20 or more | □ Always<br>□ Most of the time<br>□ Occasionally<br>□ Rarely<br>□ Never<br>□ Not applicable<br><br>What type? (Check all that apply):<br>□ Medical mask<br>□ Face shield<br>□ Gloves<br>□ Goggles/glasses<br>□ Contact gown<br>□ Impermeable gown<br>□ Coverall<br>□ Head cover<br>□ Respirator (e.g. N95 or equivalent)<br>□ Shoe covers |
| | **How many times have you been involved in the following procedures** | **What proportion of the [illegible] wearing appropriate PPE[illegible]** |

BRANY
Version Date:
05/29/2020

| | | |
|---|---|---|
| | **(in the patient room/at bedside)?** | |
| **Administration of nebulized medication** | □ None<br>□ 1-4<br>□ 5-9<br>□ 10-19<br>□ 20 or more | □ Always<br>□ Most of the time<br>□ Occasionally<br>□ Rarely<br>□ Never<br>□ Not applicable<br><br>What type? (Check all that apply):<br>□ Medical mask<br>□ Face shield<br>□ Gloves<br>□ Goggles/glasses<br>□ Contact gown<br>□ Impermeable gown<br>□ Coverall<br>□ Head cover<br>□ Respirator (e.g. N95 or equivalent)<br>□ Shoe covers |
| **Non-invasive positive pressure ventilation (CPAP/BIPAP/high flow nasal cannula)** | □ None<br>□ 1-4<br>□ 5-9<br>□ 10-19<br>□ 20 or more | □ Always<br>□ Most of the time<br>□ Occasionally<br>□ Rarely<br>□ Never<br>□ Not applicable<br><br>What type? (Check all that apply):<br>□ Medical mask<br>□ Face shield<br>□ Gloves<br>□ Goggles/glasses<br>□ Contact gown<br>□ Impermeable gown<br>□ Coverall<br>□ Head cover<br>□ Respirator (e.g. N95 or equivalent)<br>□ Shoe covers |
| | **How many times have you been** | **What proportion of the** |

BRANY
Version Date:
05/29/2020

| | **involved in the following procedures (in the patient room/at bedside)?** | **wearing appropriate PPE?** |
|---|---|---|
| **CPR involving chest compressions?** | □ None<br>□ 1-4<br>□ 5-9<br>□ 10-19<br>□ 20 or more | □ Always<br>□ Most of the time<br>□ Occasionally<br>□ Rarely<br>□ Never<br>□ Not applicable<br><br>What type? (Check all that apply):<br>□ Medical mask<br>□ Face shield<br>□ Gloves<br>□ Goggles/glasses<br>□ Contact gown<br>□ Impermeable gown<br>□ Coverall<br>□ Head cover<br>□ Respirator (e.g. N95 or equivalent)<br>□ Shoe covers |

Do you have any suggestions for the health system to improve preparedness and promote the safety of healthcare workers?
