## Supplementary Tables and Figures for "Seroepidemiology among Employees of New York City Health and Hospitals during the First Wave of the SARS-CoV-2 Epidemic"

**Supplements**

**eTable 1.** Exploratory Post-Hoc Analyses of Age group by SARS CoV-2 antibody status

**eTable 2.** Distribution of Positive SARS-CoV-2 Cases by Borough

**eFigure1.** Demographic characteristics of the study sample, H+H employee sample, and H+H workforce

**eFigure2.** Percent seropositivity by demographic characteristics of the study sample, H+H employee sample, and H+H workforce

**eTable 1. Exploratory Post-Hoc Analyses of Age group by SARS CoV-2 antibody status**

| **Age Group** | **Negative for SARS-CoV-2 IgG (n=513)** | **Positive for SARS-CoV-2 IgG (n=214)** | **Significance** | **Overall Sig** |
| --- | --- | --- | --- | --- |
|  |  |  |  | 0.38 |
| 21-34 | 126 (25%) | 42 (20%) | Ref |  |
| 35-44 | 117 (23%) | 56 (26%) | 0.15 |  |
| 45-54 | 118 (23%) | 56 (26%) | 0.15 |  |
| 55+ | 150 (30%) | 59 (28%) | 0.56 |  |

**eTable 2. Distribution of Positive SARS-CoV-2 Cases by Borough**

| **Borough** | **Study Distribution of Positive Antibody Tests (NYC residents only, n=166)^a^** | **City Distribution of Cumulative COVID Cases from 2/29/20-6/30/20^b^** | **Cumulative COVID Cases in NYC from 2/29/20-6/30/20^b^** |
| --- | --- | --- | --- |
| Queens | 34% | 30% | 74,542 |
| Brooklyn | 25% | 28% | 67,804 |
| Bronx | 20% | 22% | 53,842 |
| Manhattan | 18% | 13% | 31,204 |
| Staten Island | 3% | 7% | 17,014 |
| Overall | 100% | 100% | 244,415 |

^a^Percentages are column percentages.

^b^Data retrieved from the New York City Department of Health and Mental Hygiene’s publicly available GitHub website: <https://github.com/nychealth/coronavirus-data>

**eFigure1. Demographic characteristics of the study sample, H+H employee sample, and H+H workforce.**

A: Distribution of sex by study sample, H+H employee sample, and H+H workforce

Sex Representation

|  | Study Sample | H+H Testing Sample | H+H Workforce |
| --- | --- | --- | --- |
| Female | 74.6905% | 70.1837% | 69% |
| Male | 23.6589% | 29.7116% | 31% |

B: Distribution of race/ethnicity by study sample, H+H employee sample, and H+H workforce

Combined Race/Ethnicity Representation

|  | Study Sample | H+H Testing Sample | H+H Workforce |
| --- | --- | --- | --- |
| Asian/PI | 21.0454% | 14% | 21% |
| Hispanic | 20.0825% | 5.9% | 19% |
| Non-Hispanic Black | 17.6066% | 29.2% | 44% |
| Non-Hispanic White | 36.3136% | 22.5% | 15% |
| Other/Missing | 4.95186% | 28.5% | 1% |

Of note, the overall H+H testing sample also included American Indian (n=152), but it has been collapsed into other since it was not recorded in the study sample or overall workforce demographics. In addition, the study sample included Pacific Islander (n=5) and Multiracial (n=11) as separate categories, but they have been collapsed into Asian/Pacific Islander and Other respectively to more easily compare to the other groups.

C: Distribution of age by study sample and H+H employee sample. Age distribution data was not available for H+H workforce.

Age Representation

|  | Study Sample | H+H Testing Sample |
| --- | --- | --- |
| 18-44 | 47.0994% | 47.8% |
| 45-64 | 47.2376% | 46.7% |
| 65+ | 5.66298% | 5.5% |

**eFigure2. Percent seropositivity by demographic characteristics of the study sample, H+H employee sample, and H+H workforce.**

A: Percent seropositivity by sex

Percent Positive by Sex

|  | Study Sample | H+H Testing Sample |
| --- | --- | --- |
| Female | 30.3867% | 24.1834% |
| Male | 27.3256% | 24.0268% |

B: Percent seropositivity by race/ethnicity

Percent Positive by Race/Ethnicity

|  | Study Sample | H+H Testing Sample |
| --- | --- | --- |
| Asian/PI | 22.8758% | 20.2167% |
| Hispanic | 32.1918% | 30.6122% |
| Black | 42.1875% | 32.0172% |
| White | 25.3788% | 16.9732% |
| Other | 36.6667% | 23.4123% |

C: Percent seropositivity by race/ethnicity for Study Sample and H+H Testing Sample. Age distribution data was not available for H+H workforce.

Percent Positive by Age Group

|  | Study Sample | H+H Testing Sample |
| --- | --- | --- |
| 18-44 | 28.739% | 22.1833% |
| 45-64 | 30.4094% | 26.1819% |
| 65-74 | 26.8293% | 23.5897% |
| 75+ | 0% | 23.2877% |
